# Rapid antifouling nanocomposite coating enables highly sensitive multiplexed electrochemical detection of myocardial infarction and concussion markers

**DOI:** 10.1101/2021.06.13.21258856

**Authors:** Sanjay S. Timilsina, Nolan Durr, Mohamed Yafia, Hani Sallum, Pawan Jolly, Donald E. Ingber

**Affiliations:** Wyss Institute for Biologically Inspired Engineering, Harvard University, USA; Vascular Biology Program, Boston Children’s Hospital, and Harvard Medical School, USA; Harvard John A. Paulson School of Engineering and Applied Sciences, Harvard University, USA

**Keywords:** Antifouling, Biosensor, Electrochemical sensor, Biomarkers, Multiplexing, Myocardial Infarction, Surface chemistry, Traumatic Brain Injury

## Abstract

Here we describe an ultra-fast (< 1 min) method for coating electrochemical (EC) sensors with an anti-fouling nanocomposite layer that can be stored at room temperature for months, which provides unprecedented sensitivity and selectivity for diagnostic applications. We leveraged this method to develop a multiplexed diagnostic platform for detection of biomarkers that could potentially be used to triage patients with myocardial infarction and traumatic brain injury using only 15 µL of blood. Single-digit pg/mL sensitivity was obtained within minutes for all the biomarkers tested in unprocessed human plasma samples and whole blood, which is much faster and at least 50 times more sensitive than traditional ELISA methods, and the signal was stable enough to be measured after one week of storage. The multiplexed EC sensor platform was validated by analyzing 22 patient samples, which demonstrated excellent correlation with reported clinical values.

---

Use of an efficient and scalable anti-fouling surface chemistry is a critical requirement for commercializing reliable electrochemical (EC) biosensors for diagnostic applications. This is because many materials present in high concentrations in complex biological fluids (e.g., blood, plasma, serum, urine) generate background noise when they adsorb to the surface of EC sensors, thereby decreasing the electrical signal and specificity of the sensor which limits their commercial use.^1-5^ This has led to great interest in coating of EC sensors with anti-fouling materials, such as carbon materials, metallic nanoparticles, biopolymers, or hydrogels, as well as use of nanoporous materials.^3, 5, 6^ These materials can be applied via multiple coating methods, including physical adsorption,^7^ PEG Grafting,^8^ zwitterionic peptide self-assembled monolayer,^9^ electrochemical copolymerization,^10, 11^ drop-casting,^12^ and magnetic core-shell polymerization,^13^ but none of them are fast or simple enough to enable robust and cost-effective mass manufacturing. Moreover, some of them have reduced sensitivity due to poor conductivity. Therefore, depositing stable and reliable antifouling coatings directly on EC sensors through simple, scalable, and economical methods is extremely important to pave the way for more robust EC biosensors, which could transform the future of point-of-care (POC) diagnostics given their ability to be mass manufactured at low cost.^5^

We previously described a highly effective anti-fouling nanocomposite coating composed of denatured bovine serum albumin (BSA) cross-linked to gold nanomaterials or reduced graphene nanoparticles (rGOx) using glutaraldehyde (GA).^12, 17^ While this method was simple and greatly suppressed signal noise due to biofouling, it took 24 hours to prepare these surfaces. Here, we report an alternative version of this nanocomposite coating that is composed of BSA cross-linked to pentaamine graphene flakes that can be coated on EC sensors using localized heating in the unprecedented time of less than one minute, which allows for outstanding signal transduction between the gold electrode and the nanocomposite surface. We demonstrate the multiplexing capability and potential clinical value of these coated EC sensors by developing a multiplexed EC diagnostic for multiple blood biomarkers that can be used to triage patients with myocardial infarction (MI) or traumatic brain injury (TBI), and confirming that the values obtained within minutes using this platform with less than one drop of patient blood scale closely with those previously obtained using commercial clinical diagnostics. As this coating method is extremely fast, simple, inexpensive, and scalable, it could enable mass manufacturing of highly sensitive and selective EC sensors for diagnostic as well as industrial applications in the future.

## Ultra-fast deposition of an antifouling coating on EC sensors

To develop an efficient process flow for mass manufacturing, a nanocomposite coating method was developed using on-chip heating to facilitate rapid deposition of our previously described BSA/rGOx composite.^17^ Clean gold electrodes were heated at 85°C with the nanocomposite material for 30 sec to 5 min, followed by washing in PBS immediately or after cooling down for 10 min. This ultra-rapid cross-linking mechanism results in creation of porous three dimensional (3D) molecular networks, and we were able to detect this reaction by measuring increase in absorbance at 265-270 nm (**Supplementary Fig. S1a**).^14^

Cyclic voltammetry (CV) was used to evaluate the electrochemical performance of the different optimization steps by monitoring the oxidation and reduction peaks along with the peak-to-peak distance of potassium ferri/ferrocyanide. The CV analysis revealed a near reversible process with a peak-to-peak distance of 143 mV,^15^ with high current densities being observed when heating times were 90 sec or below (**Fig. 1a and Supplementary Fig. S1b**), and a coating time of 45 sec was used for all subsequent studies. The sharp decrease at later times may be due to excessive crosslinking of the nanocomposite on the sensor surface, leading to passivation.

**Figure 1.**
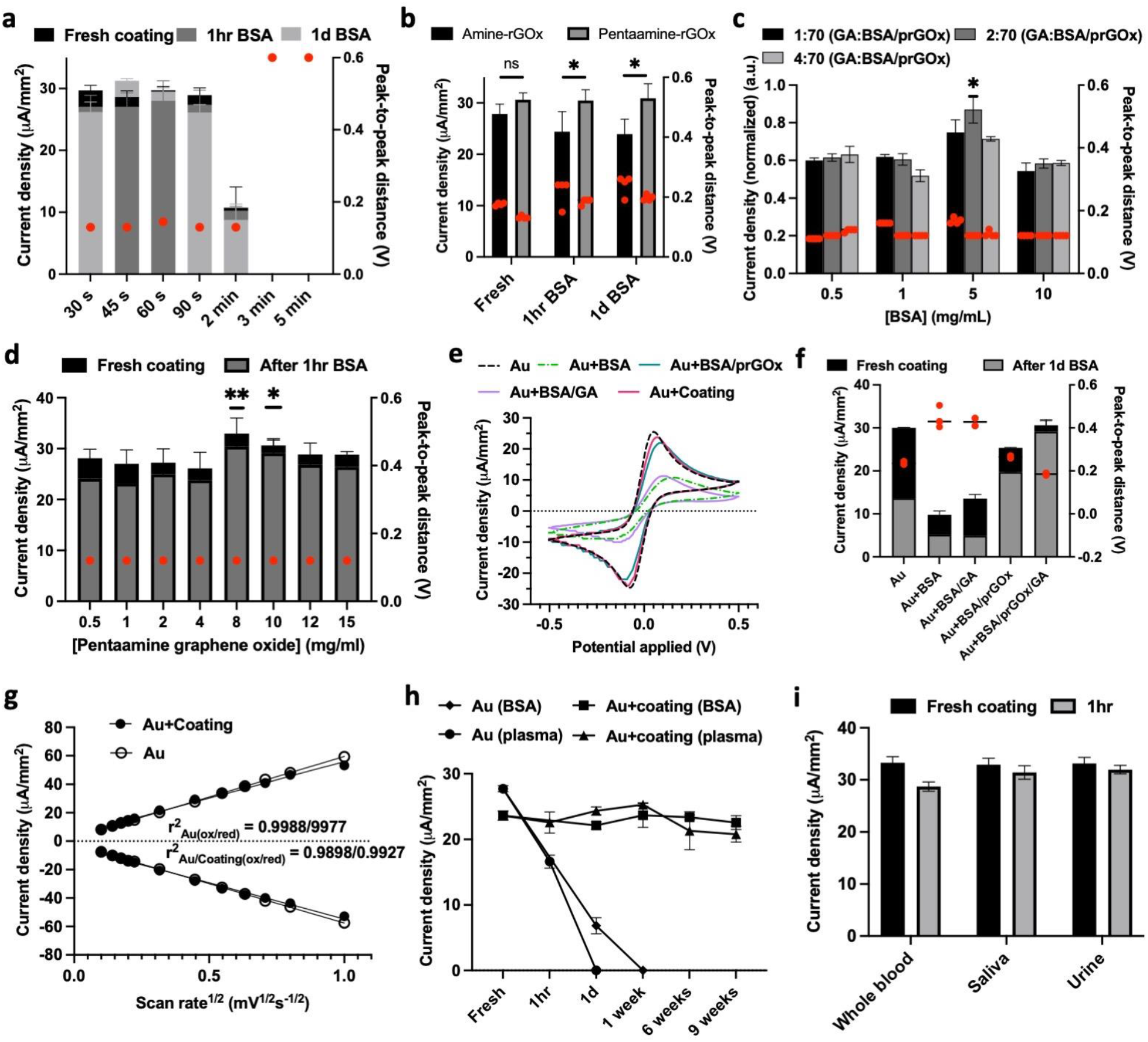
Development of antifouling nanocomposite coating method. a, Electrochemical characterization for optimization of heating time. b, Comparison of rGOx. Bars are the current density of amine functionalized GOx (black) and pentaamine functionalized GOx (grey). c, Optimization of coating mixture. Bars are the normalized values of the current density of nanocomposite with different concentration of BSA at different ratio of GA to BSA/prGOx. d, Optimization of prGOx concentrations. Bars are the current density of various nanocomposite with different concentrations of prGOx; fresh coating (black) and after 1h exposure to 1% BSA (grey). e, Typical voltammograms showing oxidation and reduction peaks of ferri-/ferrocyanide with various nanocomposite-coated electrodes. f, Electrochemical characterization of various electrode coatings. Bars are the mean current density of various nanocomposite-coated electrodes fresh (black) and after 1d exposure to 1% BSA (grey). g, Extracted oxidation/reduction peak current mean values for gold electrode (black circles) and electrode with coating (white circle) from the voltammograms shown in Supplementary Fig. S1c. h, Comparison of antifouling activity with mean value of current density recorded of bare gold electrodes and Gold electrodes with antifouling coating stored for 9 weeks at 4 °C in 1% BSA and unprocessed human plasma. i, Characterization of antifouling nanocomposite coating. Bar graph shows current density of fresh sensors (black bar) and sensors after 1h in varying biofluids (grey bar). In a, b, c, d, and f, red circles denote the final mean value of peak-to-peak distances. Statistical analysis in b, c, and d performed using unpaired t-tests: *P<0.05, **P<0.01, all two tailed.

Incorporating terminal amine groups in rGOx augments their interaction with polymer matrices and their distribution by altering their solubility and agglomeration, in addition to enhancing the physical, mechanical, thermal, and electrical stability of the composite via covalent linking of the amines by GA pyridine polymers.^16, 17^ Indeed, when we compared the effects of amine-functionalized rGOx we used previously versus pentaamine-functionalized rGOx (prGOx), we found that the prGOx maintained a higher current density and lower peak-to-peak distance when measured immediately after coating or after incubating in 1% BSA for 1h or 1 day (**Fig. 1b**). These data indicate that addition of the terminal pentaamine significantly increased the coating’s antifouling activity. Further optimization was carried out by increasing the concentration of BSA in the composite from 0.5 to 5 mg/mL (with all GA ratios), but further increases resulted in a decrease in current density, although peak-to-peak distance remained similar (**Fig. 1c**). The insulation effect at higher concentration could be due to increased cross-linking as GA can react with proteins through various mechanisms depending upon different forms it can take in solution, which is primarily developed through empirical observation.^18^ As the highest conductance was obtained with 5 mg/mL BSA and a 2:70 ratio of GA: BSA, these conditions were then used to optimize the prGOx concentration. When this method was used to coat EC sensors, the highest nanocomposite conductivity was observed at 8 mg/mL prGOx and there was no significant decrease in current even after 1h incubation in 1% BSA (**Fig. 1d**), thus confirming the excellent anti-fouling activity of the nanocomposite.

We have previously shown that gold electrodes coated with BSA/rGOx/GA maintain a higher current density (84.5%) than when coated with BSA or BSA/GA, which is likely due to electron transfer medicated by the conducting nanoparticles in the rGOx containing composite.^19^ When CV was used to compare gold electrodes that were modified with various coatings (BSA alone, BSA/GA without conducting nanoparticles, and BSA/prGOx/GA) that were exposed to 1% BSA for 24h, we found that the electrodes exhibited reduced electrochemical performance when uncoated or coated with composites without conducting nanoparticles (**Fig. 1e**). The BSA and BSA/GA coated sensors displayed a broad peak-to-peak separation (ΔEp) from this redox process up to 0.443 V, indicating limited diffusion of ferri-/ferrocyanide to the electrode surface due to fouling (**Fig. 1f**).^20^ Even after 1-day of incubation in 1% BSA, the coating with BSA/prGOx/GA maintained a very high current density of 95.1% and low ΔEp of 185 mV. Furthermore, when mass transport of potassium ferri-/ferrocyanide in the BSA/prGOx/GA-coated electrodes was assessed using CV at different scan rates, the anodic and cathodic peak currents increased with the change in scan rate (**Supplementary Fig. S1c**). This analysis also revealed excellent linear relationships for both the oxidation (r^2^ gold = 0.998, r^2^ gold-coating = 0.989) and reduction (r^2^ gold = 0.997, r^2^ gold-coating = 0.992) currents as well as the square root of scan rate (**Fig. 1g**), which indicates a surface diffusion-controlled redox electrode process.

As our long term goal is to leverage this anti-fouling coating method for creating multiplexed EC sensors for clinical diagnostics, we next explored its value for use with more complex biological fluids. The rapidly coated BSA/prGOx/GA nanocomposite showed excellent anti-fouling properties even when incubated for up to 9 weeks in either 1% BSA or unprocessed human plasma, with 95.5 and 88% of current densities being maintained, respectively, even after 9 weeks, whereas all signal was lost within 1 day when bare electrodes were exposed to these solutions (**Fig. 1h**). Importantly, there also was no significant reduction in current density when the BSA/prGOx/GA-coated sensors were exposed to whole human blood, saliva, or urine for 1h prior to analysis (**Fig. 1i**).

## Surface characterization of the antifouling coating

To better understand the enhanced electrochemical properties of the BSA/prGOx/GA coating, transmission and scanning electron microscopy (TEM and SEM, respectively) were carried out which revealed that this coating is approximately 4.5 nm thick (**Supplementary Fig. S2a**). Imaging of the BSA/prGOx/GA coated electrodes revealed a heterogenous sponge-like matrix with average pore size of 16.3 nm that occupied 11% of the total surface area, which were not apparent in bare gold or in the BSA/prGOx coating (**Fig. 2a**). This more highly porous structure explains the greater surface-diffusion controlled redox reaction observed in the BSA/prGOx/GA coated electrodes (**Fig. 1f,g**). The heterogeneity of the surface was further confirmed using atomic force microscopy (AFM) (**Fig. 2b,c and Supplementary Fig. S2b**), which depicted a mean roughness of the BSA/prGOx/GA-coated surface (2.92 nm) compared to the already rough bare gold (1.8 nm) (**Supplementary Table S1**). This increased surface roughness of the porous nanocomposite coating results in a greatly increased surface area available for interaction with biomolecules, which significantly influences the activity and concentration of surface-immobilized proteins.^21^

**Figure 2.**
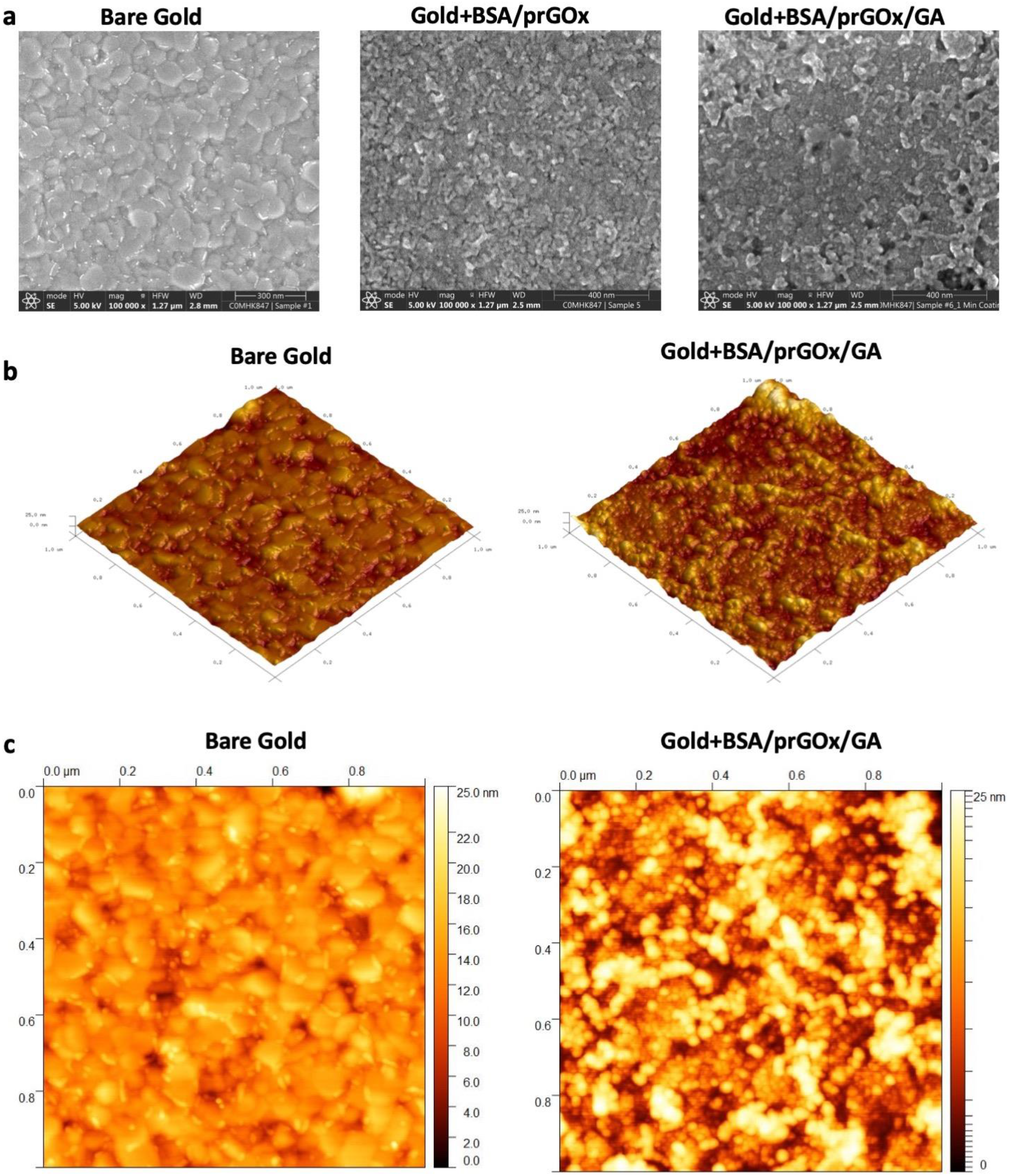
Characterization of the antifouling nanocomposite coating. a, Scanning electron micrograph of the bare Gold, Gold+BSA/prGOx, and Gold+BSA/prGOx/GA (anti-fouling coating). b, Atomic Force Microscopy (AFM) 3D images and roughness statistics for bare gold electrode and coated gold electrode. c, AFM 2D images and roughness statistics for bare gold electrode and coated gold electrode.

Evaluation of the chemical morphology of the BSA/prGOx/GA coating was also evaluated using X-ray photoelectron microscopy (XPS) and Time-of-Flight Secondary Ion Mass Spectrometry (TOF-SIMS). In the XPS spectrum, peak of C(1s) peak moved from 284 eV and focuses around 284.7 eV, corresponding to the overlap of C-C, C-H, C=C bonds. C(1s) centered peak around 288 eV with two binding energy, 287.8 (C=O, N-C=O bond) and 286.1 (C-O, C-N bond) (**Fig. 3a**), which could be assigned to overlap chemical groups such as [R-CH2-NH-(C*O)-] and [(R-CH2*-NH-(CO)-] respectively characteristics of protein and overlap (C=N, C-N and C-O bonds).^22-24^ With regards to the binding energy of the N(1s) spectrum, we observed a central peak around 400 eV corresponding to pure N(1s) peak at 399 eV along with peaks at 398.3, 398.8, 399.8, and 400.8 eV (**Fig. 3c**), which corresponds to C-N, C≡N, C=N, and NO bonds, respectively.^22, 23^ The high-resolution spectrum for the Au4f appears as two doublets located in the energy range characteristic for Au_2_O_3_ and Au (**Fig. 3d**) and O (1s) (**Fig. 3b**) around 531 eV, which is consistent with past findings.^24^ These data are consistent with successful immobilization of the BSA/prGOx/GA coating on the gold sensor electrode.^25^

**Figure 3.**
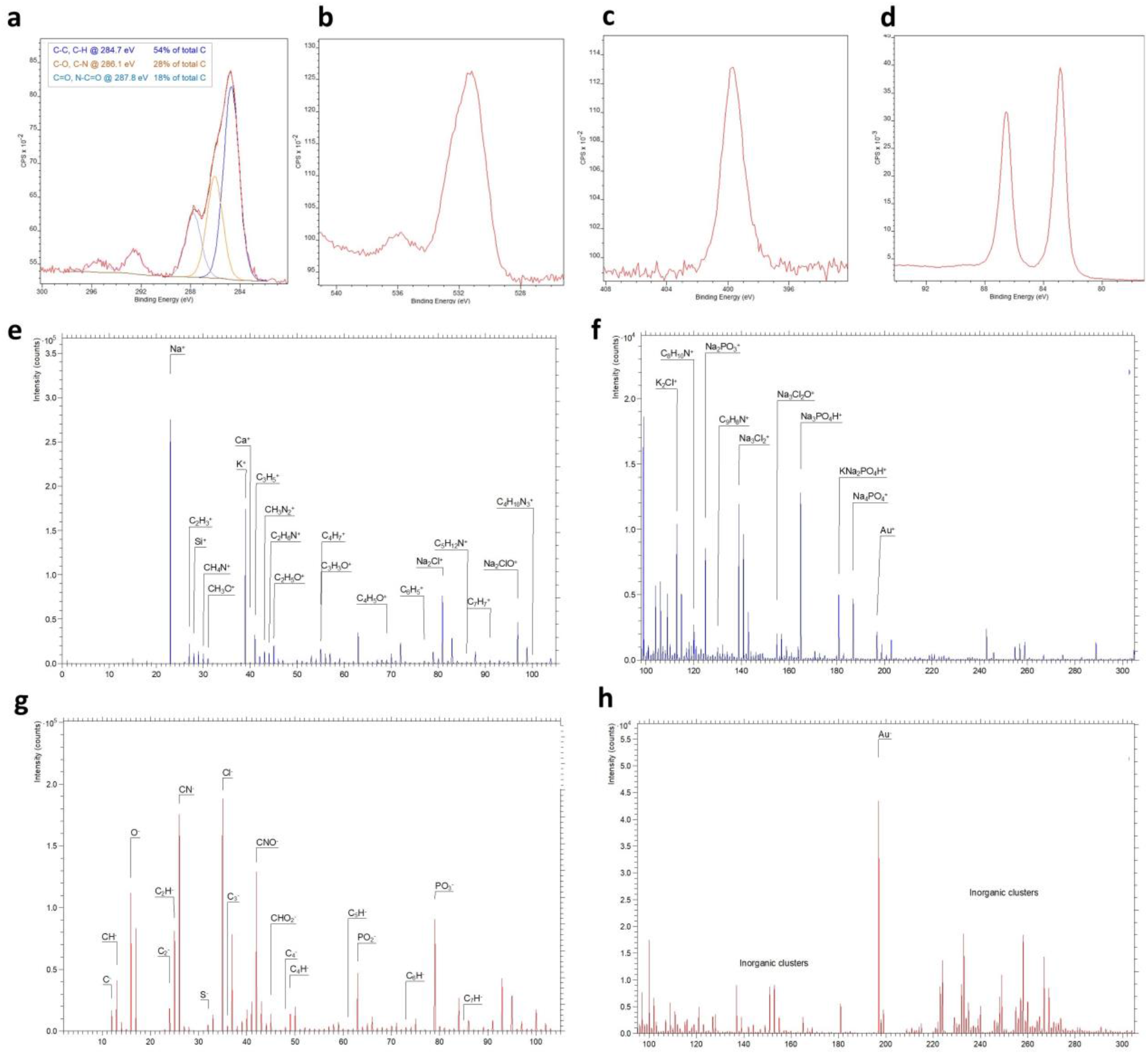
Characterization of the antifouling nanocomposite coating. X-ray Photoelectron Spectroscopy (XPS) spectrum of gold electrode with antifouling coating for C1s peak (a), O1s peak (b), N1s peak (c), and Au4f peak (d). Time-of-Flight Secondary Ion Mass Spectrometry (TOF-SIMS) spectra of gold electrode with antifouling coating for positive ions (e) and (f) and negative ions (g) and (h), where Y-axis shows the number of secondary ions detected versus X-axis of mass-to-charge (m/z) ratio of the ions.

Depth profile analysis of the nanocomposite coated gold electrode using TOF-SIMS also detected ions characteristic of BSA including CH_4_N^+^, CH_3_N ^+^, C_2_H_6_N^+^, C_4_H_5_O^+^, C_4_H_8_N^+^, C_3_H_8_NO^+^, C_5_H_12_N^+^, C_3_H_8_NO^+^, C_5_H_12_N^+^, C_4_H_10_N ^+^, C_8_H_10_N^+^, and C_9_H_8_N^+^ (**Fig. 3e,f**). Hydrocarbon species, aromatic species, and small oxygen-containing organic ions, including C^-^, CH^-^, C_2_^-^, C_2_H^-^, C_3_^-^, C_4_ ^-^, C_4_H^-^, C_5_H^-^, C_7_H^-^, CN^-^, and CNO^-^ were observed as well, as previously seen by others,^26-28^ indicating the presence of graphene oxide and BSA (**Fig. 3g,h**). The antifouling coating also increased the sensor’s hydrophilicity compared to BSA and BSA/GA (**Supplementary Fig. S3**), which can help to facilitate the binding of capture antibody.

## Ultra-sensitive analytical performance of the nanocomposite-coated EC sensor

BSA/prGOx/GA coated gold electrodes were used to create EC biosensors by immobilizing specific capture antibodies on the coating surface using 1-ethyl-3-(−3-dimethylaminopropyl) carbodiimide hydrochloride/N-hydroxysuccinimide chemistry, and detecting the antibody-bound ligand in a sandwich assay with a second biotin-labelled antibody directed against the ligand in combination with poly-streptavidin-horseradish peroxidase (HRP), and a precipitating form of tetramethylbenzidine substrate (TMB)^12^ (**Fig. 4a**). We developed a single-step assay in which sample mixed with soluble detection antibody was immediately added to the capture antibody-coated EC sensor for 30 min, which reduced the total assay time to 36 min. We then optimized the poly-streptavidin-HRP concentration (5 μg/mL), and TMB incubation time (2 min) to meet the clinical cut-off range of each biomarkers. Using this optimized assay with antibodies directed against the important MI biomarker, cardiac troponin I (cTnI), we found that current density from the adsorbed TMB was directly proportional to the cTnI concentration (0.01-10 ng/mL) and the limit of detection (LOD) was ∼9 pg/mL when spiked in human plasma (**Fig. 4b**). In contrast, when a similar assay was run in a traditional ELISA plate, the LOD was ∼67 pg/mL (**Supplementary Table S2**).

**Figure 4.**
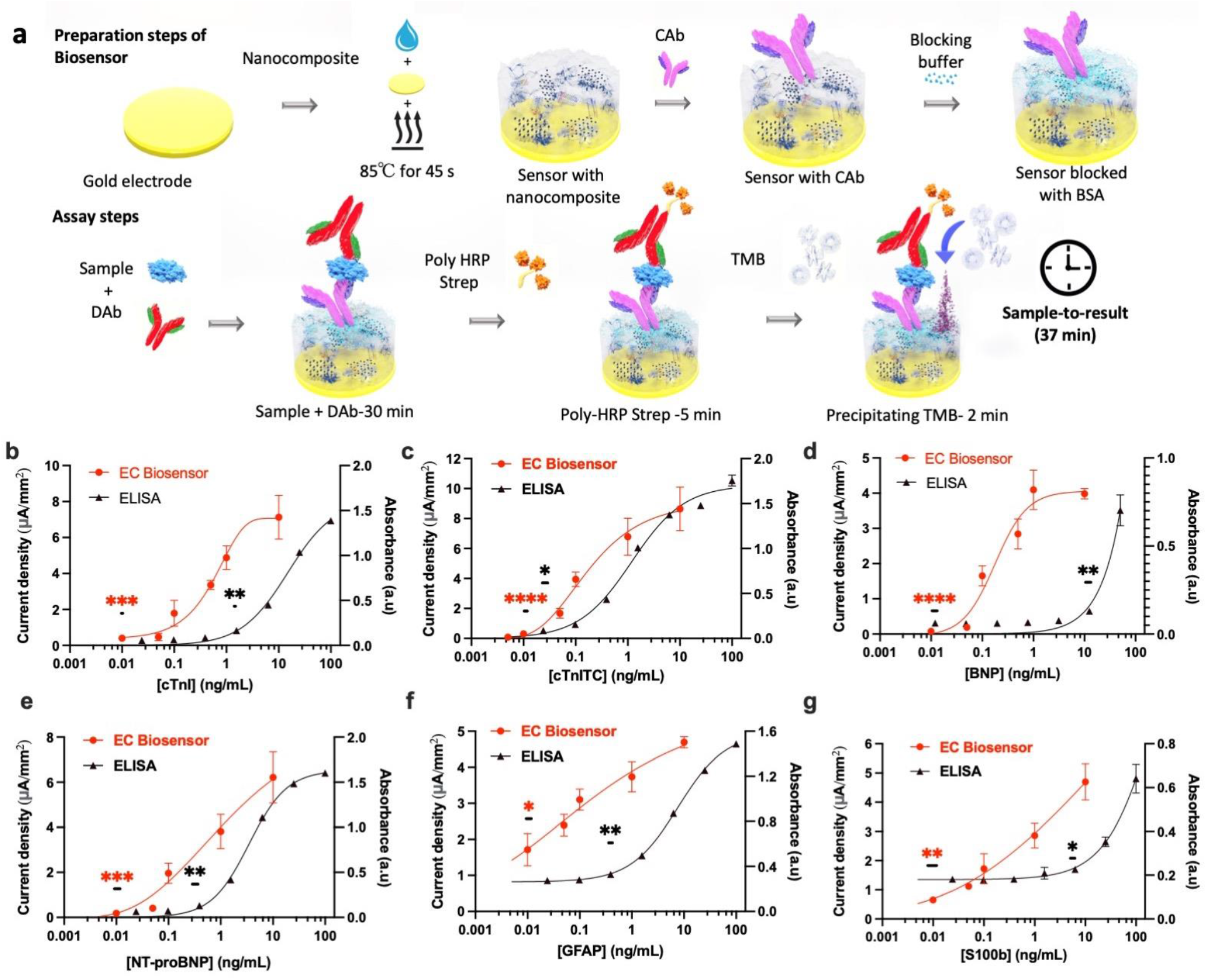
Schematic and Calibration curves for different biomarkers of Myocardial infarction and Traumatic Brain Injury using antifouling nanocomposite coated EC Biosensors and ELISA. a, Schematic for the preparation and assay steps for the EC Biosensor. b-g, Calibration curves for different biomarkers. Left y-axis shows current density for different concentrations of biomarkers run on EC biosensors (red circle) using unprocessed human plasma while right y-axis shows mean absorbance (a.u) for different concentration of biomarkers run in ELISA plate using plasma (black triangle). Different biomarkers tested include b, cardiac troponin I (cTnI); n=6, c, cardiac troponin ITC complex (cTnI-TC); n=5, d, B-type natriuretic peptide (BNP); n=4, e, N-terminal (NT)-pro hormone BNP (NT-proBNP); n=4, f, Glial fibrillary acidic protein (GFAP); n=3, and g, S100b protein; n=3. Error bars represent the s.d. of the mean; n=2 for all ELISA assay. Analysis was done using 4-Parameter Logistic (4PL) curve fitting. Lowest concentration showing significant difference against background (0 ng/mL) was determined by unpaired *t*-test (^ns^ *P > 0*.*05;* \**P* < 0.05; \*\**P* < 0.01 \*\*\**P* < 0.001; \*\*\*\**P* < 0.0001; all two tailed.

Cardiac troponins released into the bloodstream are mainly present in non-covalent ternary complex—cTnI-TC or binary cTnI-C complex—along with some free cTnI. When we attempted to detect the cTnI-TC complex in plasma, we found the LOD of the nanocomposite sensor to be much lower than in the ELISA (3 vs. 24 pg/mL, respectively)(**Fig. 4c** and **Supplementary Table S2**), and this is better than the 99th percentile cutoff value (28 pg/mL) in a healthy population used for the clinically approved cardiac troponin assay.^29, 30^ Importantly, the nanocomposite coated biosensor also showed ultra-high selectivity with no background signal for commonly found interfering molecules in biological samples, such as uric acid, dopamine, and tryptophan (**Supplementary Fig. S4**).

We extended this analysis to detect two other MI biomarkers, B-type natriuretic peptide (BNP) and its N-terminal fragment (NT-proBNP), which show variable levels in circulation according to the clinical condition and timing of measurement and, thus require a rapid and sensitive POC device for clinical use.^31^ Our nanocomposite-coated EC sensor displayed a sensitivity for BNP of 26 pg/mL, which is almost 1000-fold better than ELISA (2.8 ng/mL) (**Fig. 4d**). Likewise, the LOD for NT-proBNP in plasma was 4 pg/mL for the EC sensor compared to 65 pg/mL in the ELISA (**Fig. 4e**). Importantly, the sensitivity for EC sensor was far better than the clinical cut-off values for BNP (35 pg/mL) and NT-proBNP (125 pg/mL) that have high negative predictive values (0.94–0.98) and can be used for ruling-out heart failure according to clinical guidelines.^32^

Changes in the expression of Glial fibrillary acidic protein (GFAP) and S100b markers have been shown to correlate with injury magnitude, survivability, and neurologic outcome of TBI.^33^ As with the detection of the MI biomarkers, we found that the coated EC sensor was a much more sensitive detector of these TBI biomarkers than the standard ELISA assay (LOD of 5 vs. 59 pg/mL for GFAP and 1 vs. 957 pg/mL for S100b, respectively) (**Fig. 4f,g**), and these sensitivities were again better than the current clinical cut-off values (230 pg/mL for GFAP and 470 pg/mL for S100b).^33^ Moreover, when calibration curves for the cTnI-TC, GFAP, and S100b assays were carried out with unprocessed human whole blood, the LODs were 22, 2, and 13 pg/ml, respectively (**Supplementary Fig. S5**). Finally, to reduce the overall assay time, enhance analyte transport, and increase binding kinetics, a microfluidic device was developed with 6 integrated nanocomposite-coated EC sensors for multiplexed parallel testing of six sensors where LOD of 14 pg/mL and 27 pg/mL was obtained for cTnI-TC and GFAP, respectively, within 15 min (**Supplementary Fig. S6**).

## Stability of the nanocomposite-coated EC biosensors

To explore the future utility of this nanocomposite coating method, we compared the rapid (45 sec) coating method with the previously published 24h coating method^12^, using the NT-proBNP assay. We found that the rapid coating is as efficient as the longer method (**Fig. 5a**). Moreover, the TMB that precipitates on the sensor as a result of ligand detection (**Fig. 4a**) is highly stable and produces a similar current density after storage for up to 8 days at room temperature using the GFAP assay (**Fig. 5b**). To explore the robustness of the nanocomposite coating solution, it was stored at 4°C versus room temperature for up to 20 weeks. The stored sensors were then rapidly coated with nanocomposite solution and a GFAP assay was carried out. These studies revealed that >85% of the current response was maintained when the solution was stored at 4°C (**Fig. 5c**) or room temperature (**Fig. 5d**) compared to the fresh coating.

**Figure 5.**
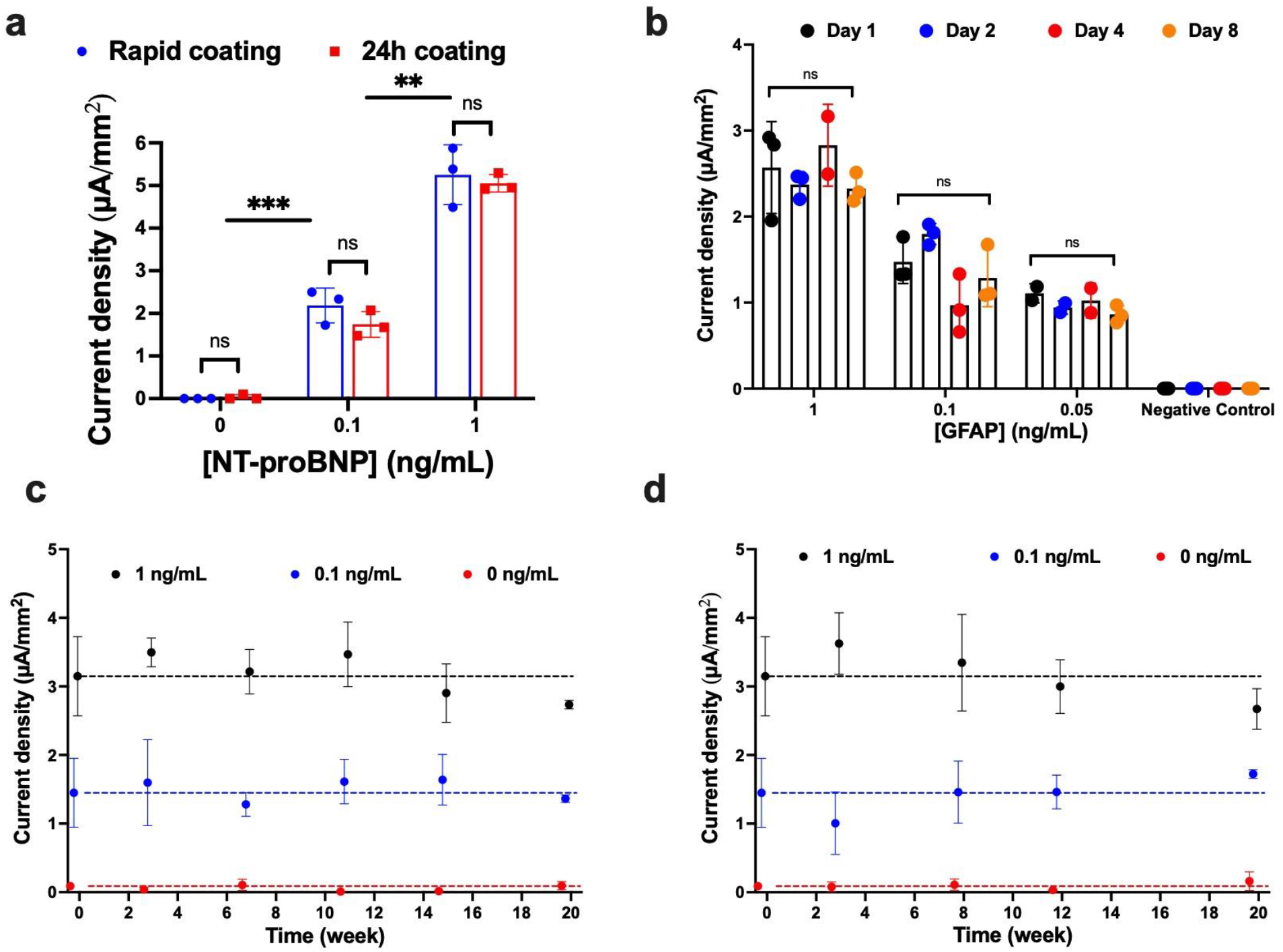
Characterization and stability of antifouling nanocomposite and precipitated TMB. a, Assay of NT-proBNP comparing the mean current density for different concentrations of NT-proBNP done on EC-Biosensor with rapid coating (blue) and 24h coating (red). b, Stability of precipitated TMB for detection of GFAP. Bars graph shows the mean current density for GFAP measured just after the assay, day 1 (black dot), day 2 (blue dot), day 4 (red dot), and day 8 (orange dot); n=3. Stability of coating solution stored at 4 degrees (c) and room temperature (d) for the assay of GFAP on EC-Biosensor coated with nanocomposite stored for 20 weeks; n=3. Error bars represent the s.d. of the mean. In a, significance was determined by multiple/unpaired t-test (ns P > 0.05; *P < 0.05; **P < 0.01 ***P < 0.001; ****P < 0.0001; all two-tailed. In b, Two-way ANOVA was used to observe a significant source of variation, ns P > 0.05.

## Multiplexed electrochemical detection of MI & TBI markers

Importantly, the high sensitivity and selectivity of the nanocomposite coated EC sensors made it possible to create a high density multiplexed sensor array and to carry out simultaneous detection of multiple clinically relevant MI and TBI biomarkers (cTnl-TC, S100b, NT-proBNP, and GFAP) by individually functionalizing each sensor’s electrode with specific capture antibodies (**Fig. 6a**). Using this multiplexed EC sensor device, we were able to specifically detect each ligand individually, and we did not detect any cross-reactivity with capture or detection antibodies (**Fig. 6b**).

**Figure 6.**
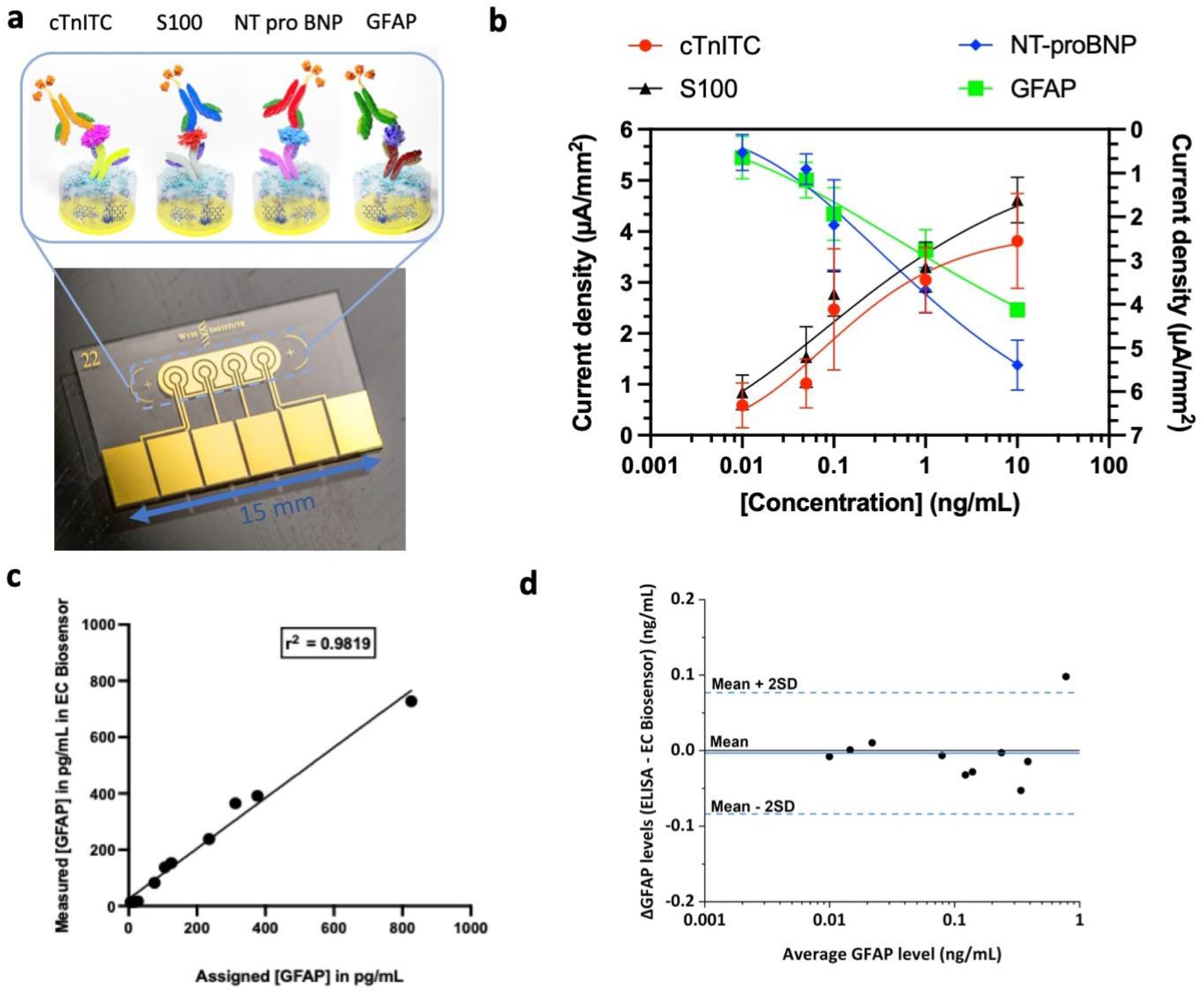
Multiplexed detection and Clinical validation for different biomarkers of MI and TBI on EC-Biosensors. a, Schematic for multiplexed detection on the EC-Biosensor showing detection of four biomarkers in a single EC-Biosensor. b, Calibration curve for multiplex detection of increasing concentration of cTnITC and S-100b (left y-axis) and decreasing concentration of GFAP and NT-proBNP (right y-axis) using plasma sample; n=3. c, Validation of the EC-Biosensor using standard ELISA assay for the detection GFAP with n=3 for each clinical sample. d, Bland-Altman plot for validation of EC-Biosensor using clinical sample for GFAP. Error bars represent the s.d. of the mean.

Finally, we tested the multiplexed sensor platform by attempting to detect cTnI-TC and GFAP in 22 patient plasma samples and compared the results to those obtained using the conventional ELISA. The linear regression analysis confirmed that there was an excellent correlation between the EC sensor and the ELISA (r^2^ = 0.9848 for cTnI-TC and 0.9819 for GFAP) (**Fig. 6c** and **Supplementary Fig. S7a)**. There also was excellent agreement between the two methods in the lower concentration region (<1 ng/mL) when analyzed using a Bland-Altman plot (**Fig. 6d and Supplementary Fig. S7b)**.

## Discussion

The clinical use of EC sensors has been limited because they suffer from surface fouling when used with biological samples. Antifouling methods have been developed^1, 3, 5, 6^; however, most have not been validated using clinical samples in complex biological fluids and they require complex manufacturing strategies that are often not consistent with mass manufacturing required for commercial translation. Here, we describe a rapid, versatile, robust, and low-cost method for coating sensors with a highly efficient antifouling nanocomposite coating accomplished using a simple method involving localized heat-induced formation of the coating that occurs in the unprecedented time of less than one minute (at least 3 orders of magnitude faster than methods previously reported in the literature). The nanocomposite provides enhanced antifouling properties while retaining over 95% of electrode conductivity. When fabrication conditions were optimized, gold electrodes retained over 88% of their sensitivity for up to 9 weeks of incubation in unprocessed human plasma, and could be stored at room temperature for at least 20 weeks with 85% current response demonstrating their long-term stability. While this antifouling nanocomposite coating has not been commercialized, and thus, only has been tested under laboratory conditions, we demonstrated its ability to detect clinically relevant biomarkers in patient-derived biological fluids with higher sensitivity and specificity than commercial ELISA assays. Furthermore, the simple and rapid antifouling process used to accomplish this could easily be translated into a reel-to-reel mass manufacturing process and thus, it has enormous potential for commercial development of a marketable device with reliable functioning.^6^

The porous BSA backbone of the matrix prevents non-specific protein adsorption but allows diffusion of electro-active soluble species. The employment of prGOx, which is highly conductive, accelerates electron transfer and enhances the transduction properties. The rapid antifouling coating method combined with extensive assay development helped to significantly improve the performance of gold electrode based EC sensors, as demonstrated by the increased sensitivity and wide dynamic and linear range of the EC sensors as compared to traditional ELISA assays. These EC sensors with this antifouling coating demonstrate excellent anti-fouling properties shown by the coating in complex biological samples, such as plasma, whole blood, saliva, and urine; they can be multiplexed and still retain high sensitivity and specificity; because they use an EC readout, they can be carried out with minimal training; and they also showed excellent correlation with reported values in clinical samples. Thus, this approach has huge potential for commercial POC diagnostic applications, for example, to combat future crises that may be similar to the current global coronavirus pandemic by detecting viral antigens and host antibodies in the field. The stability of the precipitated TMB for over a week provides additional flexibility that could also enable shipping sensors collected from large populations of patients to a central laboratory when POC detection is not desired.

In summary, the development of a simple and rapid method for coating EC sensors with a highly effective antifouling coating using methods that are scalable for mass manufacturing opens up an entirely new path for development of multiplexed EC diagnostics for POC applications. Our proof-of-concept studies demonstrated this for detection of multiple clinical biomarkers for MI and TBI using only a drop of blood, and the results were as good as or better than clinical diagnostics used for this application currently. As this EC sensor system enables rapid diagnosis with only minimal sample handling and users skills, it could be useful for diagnosis at home, as well as in pharmacies, ambulances doctor’s offices, and hospitals. While we demonstrated biosensing using detection of labels (TMB), other EC approaches, such as open circuit potential impedance spectroscopy, could be pursued to build a label-free sensing platform in the future. By replacing antibodies with other type of detection ligands (e.g., aptamers, DNA, RNA, CRISPR), even more advanced real-time biosensing could be developed.

## METHODS

### Anti-fouling nanocomposite preparation

The gold electrode chips custom fabricated using a standard photolithography process and purchased from Telic Company were cleaned as described previously.^17^ The BSA/prGOx/GA anti-fouling nanocomposite was prepared by mixing 8 mg/mL of prGOx (Millipore Sigma, no. 806579) with 5 mg/mL (IgG-Free, Protease-Free, Bovine Serum Albumin) (Jackson ImmunoResearch, no. 001-000-162) in 10 mM phosphate-buffered saline solution (PBS, pH 7.4) (Sigma Aldrich, USA, no. D8537). A similar method was used to prepare the coating with amine-functionalized reduced graphene oxide (Sigma Aldrich, USA, no. 805432). The solution was sonicated in a tip sonicator for 30 min using 1 s on/off cycles at 50% amplitude, 125 W and 20 kHz (Bransonic, CPX 3800) followed by heating (Labnet, no. D1200) at 105 °C for 5 min to denature the protein. The resulting opaque black mixture was centrifuged at 16.1 relative centrifugal force for 15 min to remove the excess aggregates. The semi-transparent nanocomposite supernatant solution was then mixed with 70% glutaraldehyde (Sigma Aldrich, USA, no. G7776) for crosslinking in the ratio of 70:2.

### Development of coating strategy of anti-fouling nanocomposite

To develop the best coating strategy of the anti-fouling nanocomposite to the gold electrodes of the chips; cleaned and plasma-treated chips were first kept over a hot plate for 2 min to let the temperature equilibrate to 85 °C. 70 μL of anti-fouling nanocomposite as described above was then drop cast to each chip and incubated for 30s-5 minutes. The chips were then washed by dipping in PBS immediately or after cooling down for 10 min followed by washing at 400 rpm for 10 min. The chips were then exposed to 1M ethanolamine (Sigma Aldrich, USA, no. E9508) in PBS to quench the unreacted glutaraldehyde groups before electrochemical analysis. For optimization of BSA-glutaraldehyde ratio of the anti-fouling coating, nanocomposite solution was prepared with different concentrations of BSA ranging from 0.5 mg/mL to 10 mg/mL with 3 different ratios of BSA & GA (1:70, 2:70, and 4:70). The chips were coated with these solutions for 45s at 85 °C followed by washing and electrochemical analysis. To further develop the nanocomposite that can retain the maximum signal and have the best anti-fouling properties, nanocomposite was prepared with 5 mg/mL BSA with different concentration of prGOx (0.5, 1, 2, 4, 8, 10, 12, 15 mg/mL) with GA: BSA/prGOx of 2:70. Electrochemical analysis was done after quenching the reaction with 1M ethanolamine for 30 min, followed by 1h incubation in 1% BSA.

### Electrochemical characterization of anti-fouling nanocomposite

All the electrochemical characterization and assay were carried out on the chip containing four working gold electrodes, a common pseudo-reference gold electrode, and a common gold counter-electrode connected to a potentiostat (Autolab PGSTAT128N, Metrohm; VSP, Bio-Logic) through an in-house built connector device. The conductive anti-fouling nanocomposite was electrochemically characterized by cyclic voltammograms (CV) in a redox aqueous solution of PBS containing 5 mM [Fe(CN)_6_]^3-/4-^ at a scan rate of 200 mV/s between -0.5 to 0.5V. For anti-fouling properties chips were incubated with plasma (Innovative Research, USA, no. IPLASCOV2P100UL), whole blood (Innovative Research, USA, no. IWB1NAH10ML), urine (drug/alcohol-free, LEE Biosolutions, USA, no.991-03-DF-50), and saliva (avantor, USA, no. 102768-970) for different time before electrochemical measurement. In addition, to characterize the reversibility of the system, bare gold electrodes and gold electrodes covered with nanocomposite were analyzed by CV in PBS containing 5 mM [Fe(CN)_6_]^3-/4-^ by increasing the scan rate from 10 to 1000 mV/s between -0.5 to 0.5V.

### Spectroscopic/Microscopic study of gold electrode/anti-fouling nanocomposite

The nanocomposites along with intermediate components were characterized by UV spectroscopy (Nanodrop 2000c, Thermo Scientific) after the addition of every component, to elucidate changes in the absorbance bands of the peptide backbone or the aromatic rings due to the crosslinking of BSA and prGOx with GA. Topographic characterization for the nanocomposite and other coatings over the sensor was carried out by SEM (Verios G4 XHR, ThermoFisher Scientific). The samples were first coated with a thin layer of approximately 6 nm Cr by sputtering (K755X EM Technologies LTD.). Imaging was carried out by detection of secondary electrons using an in-lens detector at an accelerating voltage of 5 kV. Porosity was calculated from SEM images using the method described previously.^12^ For TEM characterization, samples were prepared using the in-situ FIB lift-out technique on an FEI Helios 650 Dual Beam FIB/SEM. The samples were capped with sputtered Ir, protective carbon and e-Pt/I-Pt before milling. To obtain better contrast of the coating one set of TEM was done without Ir. The TEM lamella thickness was ∼100nm. The samples were imaged with an FEI Tecnai Talos FEG/TEM operated at 200kV in bright-field (BF) TEM mode and high-resolution (HR) TEM mode. Similarly, AFM images were collected using a Dimension Icon AFM instrument (Bruker, Santa Barbara, California, USA). The instrument was calibrated against a NIST traceable standard. Soft Tapping Mode was used as analysis mode with OTESPA-R3 (Bruker) as AFM probe. 1^st^ order flattening was used for data post-processing. One 1 μm x 1 μm area was imaged near the center of each sample. AFM 2D and 3D height images were obtained along with the roughness measurements of gold and gold/coating. The topography differences of these images are presented in colors where the brown is low and the white is high. The z ranges are noted on the vertical scale bar on the right side of the images. X-ray Photoelectron Spectroscopy (XPS) was used to determine semi-quantitative atomic composition and chemistry using PHI Quantum 2000 instrument. Monochromated Alka_α_ 1486.6eV was used as X-ray source with an acceptance angle of ±23°, take-off angle of 45°, and analysis area of 1400 μm × 300 μm. TOF-SIMS was performed using IONTOF TOF-SIMS 5 instrument and data were obtained using a liquid metal ion gun (LMIG) primary ion source. Both surface spectrum and depth profile were acquired from the sample. Ar-cluster was used as ion source with Ion beam potential of 2.5 keV on an area of 500 μm × 500 μm for sputtering. Likewise, Bi_3_^+^ was used as ion source with Ion beam potential of 30 keV on an area of 200 μm × 200 μm for analysis. The data are presented as mass spectra which are displayed as the number of secondary ions detected (Y-axis) versus the mass-to-charge (m/z) ratio of the ions (X-axis). The ion counts are shown on linear intensity scales, and probable empirical formulae for several peaks are identified in the figures.

### Conjugation of Capture Antibodies

After coating the electrodes with nanocomposite coating, chips were washed in PBS by agitation (400 rpm) for 10 min and dried with a slide spinner (Millipore Sigma, no. 674664). 1-ethyl-3-(3-dimethylaminopropyl) carbodiimide hydrochloride (EDC, Thermo Fisher Scientific, no. 22980) and N-hydroxysuccinimide (NHS, Sigma Aldrich, no. 130672) were dissolved in 50 mM MES (2-(N-morpholino)ethanesulfonic acid) Buffer (pH 6.2) at 400 mM and 200 mM, respectively and deposited on nanocomposite covered gold chips for 30 min at room temperature in dark. After surface activation with EDC/NHS, chips were quickly rinsed with Milli Q water and dried with compressed air. Thereafter, spotting of capture antibody on top of the working electrode area using Xtend capillary microarray Pin (LabNEXT, no. 007-350) was performed. The fourth working electrode was always spotted with 5 mg/mL BSA as a negative control unless mentioned otherwise. The spotted chips were stored overnight at 4 °C in a humidity chamber. After conjugation, chips were washed with PBS and quenched with 15 μL of 1M ethanolamine (SigmaAldrich, no. E9508) for 30 min and blocked with 10 μL of 2.5% BSA in PBS for 1 hour.

### Detection of Biomarkers using the EC-Biosensor

Detection of all the biomarkers was done on the chip using the optimized conditions. Three working electrodes were spotted with a capture antibody concentration of 500 μg/mL diluted in PBS [anti-cTnI (abcam, no. ab243982), anti-cTnI-TC (Advanced ImmunoChemical, no. 2-TIC-rc), anti-NT-proBNP (Medix Biochemica, no. 100521), anti-BNP (HyTest, no. 50E1cc), Anti-GFAP (HyTest, no. GFAP83cc), and anti-S100b (HyTest, no. 8B10cc)]. Antigens were spiked into the plasma samples at different concentrations ranging from 10 pg/mL to 10 ng/mL and mixed with optimized biotin conjugated detection antibody in the ratio of 9:1 and 15 μL was added to each chip. Antigen includes [cTnI (Medix Biochemica, no. 610102), cTnI-TC complex (HyTest, no. 8T62), NT-proBNP (Medix Biochemica, no. 610090), BNP-32 (Bachem, no. 4095916), GFAP (HyTest, no. 8G45), and S100b (HyTest, no. 8S9h)], respectively. Conjugation of biotin to detection antibody was done using Biotin Conjugation Kit (Fast, Type A) - Lightning-Link® (abcam, USA, no. ab201795) using manufacturer’s protocol. Detection antibody used include anti-cTnI (abcam, no. ab243982), anti-cTnI-TC (Advanced ImmunoChemical, USA no. 2-TC), anti-NT-proBNP (Medix Biochemica, no. 100712), anti-BNP (HyTest, no. 24C5cc), anti-GFAB (HyTest, no. GFAP81cc), and anti S-100b (HyTest, no. 6G1cc), respectively. Mixture of sample-detection antibody was incubated with agitation at 400 rpm for 30 min followed by washing with PBST (PBS with 0.05 % Tween 20 (Sigma Aldrich, no. P9416)). 5 μg/mL of Poly-HRP-Streptavidin (Thermo Fisher Scientific, no. N200) diluted in 0.1% BSA in PBST was added for 5 min. Finally, precipitating 3,3′,5,5′-Tetramethylbenzidine (TMB, Sigma-Aldrich, USA, no.T9455) was added to chips and incubated for 2 min before washing and reading the chips. Finally, measurement was performed in PBST using a potentiostat by a CV with a scan rate of 1 V/s between -0.5 and 0.5 V vs on-chip integrated gold quasi reference electrode. Peak height was calculated using Nova 1.11 software for data analysis. Clinical sample validation was performed using the same optimized conditions for cTnI-TC and GFAP. All the experiments ran on the EC-Biosensor had one electrode modified with BSA instead of the capture antibody for the on-chip negative control. No current was observed, signifying that the nanocomposite coating had excellent antifouling properties, and the precipitation of TMB is highly localized without cross-contamination to the neighboring electrodes.

### Detection of Biomarkers using Standard colorimetric ELISA

For standard ELISA of the biomarkers, 100 μL of 1 μg/mL capture antibody prepared in carbonate-bicarbonate buffer at pH 9.2 was added to Nunc™ MaxiSorp™ ELISA plates (BioLegend, no. 423501) and incubated overnight at 4 °C. The plates were washed 3 times with 200 μL of PBST followed by the addition of 200 μL of 2.5% BSA for 1h. After washing the plates 100 μL of different concentrations of the biomarkers in plasma were added and incubated at 400 rpm for 1h. After washing the plates biotinylated detection antibody (0.5 μg/mL for cTnI, NT-proBNP, cTnITC, and GFAP and 1 μg/mL for BNP and S100b) was added for 1h before washing and adding 100 μL of Streptavidin-HRP (1:200 dilution in 0.1 % BSA). The plate was washed and 150 μL of turbo TMB (Thermo Scientific, no. 34022) was added for 20 min followed by the addition of 150 μL of Stop solution to stop the reaction. The plate was immediately read using a microplate reader at 450 nm. For Clinical samples of cTnI-TC, samples were diluted 1:1 in 1% BSA. For Clinical Samples of GFAP, a magnetic bead-based assay from ThermoFisher Scientific was used following the protocol mentioned in the user guide, and measurement was done using Bio-Plex 3D Suspension Array System. Clinical samples received from Discovery Life Sciences in dry ice were aliquoted and stored at -80 °C until further use. All samples were collected under the approval of the Institutional Review Board for Harvard Human Research Protection Program (IRB21-0024).

### Multiplexed detection of MI & TBI biomarkers using the EC-Biosensor

Specificity of Antigen and Antibody was performed before doing multiplexed detection. For Multiplexed detection, four different capture antibodies (anti-cTnITC, anti-S100b, anti-NT-proBNP, and anti-GFAP) was spotted on four different electrodes of the chip at 500 μg/mL. Increasing concentration of cTnITC and S100b and decreasing concentration of GFAP and NT-proBNP (0.01 cTnITC & S100b + 10 GFAP & NT-proBNP; 0.05 cTnITC & S100b +1 GFAP & NT-proBNP; 0.1 cTnITC & S100b + 0.1 GFAP & NT-proBNP; 1 cTnITC & S100b + 0.05 GFAP & NT-proBNP; 10 cTnITC & S100b + 0.01 GFAP & NT-proBNP) was mixed with all four biotinylated detection antibody and incubated for 30 min. Chips were washed and Poly-HRP-Streptavidin was added for 5 min followed by TMB for 2 min before washing and reading the chips.

### Rapid electrochemical assay with microfluidic integration of the EC-Biosensor

The microfluidic chip has several inlet and outlet ports and is covered with a special arrangement of cover and adhesive tapes to attach the sensor on top of the microfluidic chip. These tapes also define the flow channel between the sensor and the microfluidic chip (**Supplementary Fig. S6 a**,**b**). The microfluidic chips were fabricated via 3D printing (Tiger APEX pro XHD 3D printer, Tiger 3D & Romanoff Int. Amityville, NY, USA). A clear photopolymer resin with low viscosity was used to fabricate the microfluidic chip (Tiger3D Clear 78-5011-L, Tiger 3D & Romanoff Int. Amityville, NY, USA). For post-processing, chips were cleaned in an isopropanol bath and cured under UV for 5 min (Form cure FH-CU-01, Formlabs Inc., Sommerville, MA, USA). A microfluidic cover tape was used to seal the chips (3M™ Microfluidic Diagnostic Tape 9795R, 3M Medical specialties, St. Paul, MN, USA). A double-sided adhesive tape was used to bond the sensor to the microfluidic chip and to pattern the sidewalls of the microfluidic channel sandwiched between them (ARseal™90880 Polypropylene Double-Sided Adhesive Tape, Adhesive research Inc., Glen Rock, PA, USA). The flow was initiated by peristaltic microfluidic pumps (RP-QII, Tagasako, Nagoya, Japan) driven by H-bridge brushed motor drivers (Pololu, Las Vegas, NV, USA). The pumps were controlled by a microcontroller (Arduino Uno, Arduino, Turin, Italy) via pulse density modulation to allow for high flow resolution at low pump speeds. The pump system was commanded through an LCD/keypad interface (Adafruit, New York, NY, USA). To perform an assay on the microfluidic platform, the sample mixed with detection antibody (40 µl) was added to inlet wells at the flow rate of 5 µl/min for 8 min. An optimized condition of detection antibody as discussed earlier was used. 25 µl of strep-HRP was then added at 5 µl/min for 5 min. Finally, 20 µl of TMB was added at 20 µl/min for 1 min and incubated under static incubation for 2min.

## Data Availability

The main data supporting the results in this study are available within the paper and its Supplementary Information. All raw and processed data generated in this work, including the representative images provided in the manuscript, are available from the corresponding authors on reasonable request.

## Data availability

The main data supporting the results in this study are available within the paper and it’s Supplementary Information. All raw and processed images/data generated in this work, including the representative images provided in the manuscript, are available from the corresponding authors on reasonable request.

## Acknowledgments

We thank Eurofins Scientific for optical characterization. We acknowledge research funding from the Wyss Institute for Biologically Inspired Engineering at Harvard University. M.Y. acknowledges Fonds de recherche du Québec nature et technologie (FRQNT) postdoctoral fellowship #260284.

## Authors Contributions

S.S.T., N.D., P.J. and D.E.I. conceived this study. Experiments were carried out by S.S.T., N.D., M.Y., P.J. under the guidance of D.E.I. Data analysis was carried out by S.S.T, N.D, and P.J. M.Y. developed the microfluidic chip cartridge and assisted with the microfluidic detection assay. H.S. developed the automated peristaltic pump system to perform the microfluidics. P.J. conceptualized the idea, coordinated the experiments, and managed the project progress. All authors discussed the results and contributed to writing the manuscript.

## Ethics declarations

### Supplementary Information is available for this paper

Supplementary Information includes characterization of antifouling nanocomposite coating, further characterization of sensors (AFM and TEM), contact angles of various sensors, typical cyclic voltammogram for cross-reactivity study, calibration curve of biomarkers using whole blood, microfluidic design and assay, clinical validation of EC sensors, table with roughness result for AFM, and table with comparison of sensitivity and Kd values for detection of MI and TBI biomarkers.

## REFERENCES

1. Hanssen, B.L., Siraj, S. & Wong, D.K. Recent strategies to minimise fouling in electrochemical detection systems. Reviews in Analytical Chemistry 35, 1–28 (2016).

2. Jolly, P. et al.. Label-free impedimetric aptasensor with antifouling surface chemistry: A prostate specific antigen case study. Sensors and Actuators B: Chemical 209, 306–312 (2015).

3. Jiang, C. et al.. Antifouling strategies for selective in vitro and in vivo sensing. Chemical reviews 120, 3852–3889 (2020).

4. Russo, M.J. et al.. Antifouling Strategies for Electrochemical Biosensing: Mechanisms and Performance toward Point of Care Based Diagnostic Applications. ACS sensors (2021).

5. Lin, P.-H. & Li, B.-R. Antifouling strategies in advanced electrochemical sensors and biosensors. Analyst 145, 1110–1120 (2020).

6. Campuzano, S., Pedrero, M., Yáñez-Sedeño, P. & Pingarrón, J.M. Antifouling (bio) materials for electrochemical (bio) sensing. International journal of molecular sciences 20, 423 (2019).

7. Ruiz-Valdepeñas Montiel, V.c. et al. Delayed sensor activation based on transient coatings: Biofouling protection in complex biofluids. Journal of the American Chemical Society 140, 14050–14053 (2018).

8. Hui, N., Sun, X., Niu, S. & Luo, X. PEGylated polyaniline nanofibers: antifouling and conducting biomaterial for electrochemical DNA sensing. ACS applied materials & interfaces 9, 2914–2923 (2017).

9. Cui, M., Wang, Y., Wang, H., Wu, Y. & Luo, X. A label-free electrochemical DNA biosensor for breast cancer marker BRCA1 based on self-assembled antifouling peptide monolayer. Sensors and Actuators B: Chemical 244, 742–749 (2017).

10. Yang, L., Wang, H., Lü, H. & Hui, N. Phytic acid functionalized antifouling conducting polymer hydrogel for electrochemical detection of microRNA. Analytica Chimica Acta 1124, 104–112 (2020).

11. Hu, Y. et al.. Antifouling zwitterionic coating via electrochemically mediated atom transfer radical polymerization on enzyme-based glucose sensors for long-time stability in 37 C serum. Langmuir 32, 11763–11770 (2016).

12. Del Río, J.S., Henry, O.Y., Jolly, P. & Ingber, D.E. An antifouling coating that enables affinity-based electrochemical biosensing in complex biological fluids. Nature nanotechnology 14, 1143–1149 (2019).

13. Lv, S., Sheng, J., Zhao, S., Liu, M. & Chen, L. The detection of brucellosis antibody in whole serum based on the low-fouling electrochemical immunosensor fabricated with magnetic Fe3O4@ Au@ PEG@ HA nanoparticles. Biosensors and Bioelectronics 117, 138–144 (2018).

14. Cheung, D.T. & Nimni, M.E. Mechanism of crosslinking of proteins by glutaraldehyde I: reaction with model compounds. Connective tissue research 10, 187–199 (1982).

15. Zanello, P., Nervi, C. & De Biani, F.F. Inorganic electrochemistry: theory, practice and application. (Royal Society of Chemistry, 2019).

16. Saravanan, N. et al.. Graphene and modified graphene-based polymer nanocomposites– a review. Journal of Reinforced Plastics and Composites 33, 1158–1170 (2014).

17. Zupančič, U., Jolly, P., Estrela, P., Moschou, D. & Ingber, D.E. Graphene Enabled Low-Noise Surface Chemistry for Multiplexed Sepsis Biomarker Detection in Whole Blood. Advanced Functional Materials, 2010638 (2021).

18. Migneault, I., Dartiguenave, C., Bertrand, M.J. & Waldron, K.C. Glutaraldehyde: behavior in aqueous solution, reaction with proteins, and application to enzyme crosslinking. Biotechniques 37, 790–802 (2004).

19. Gooding, J.J., Alam, M.T., Barfidokht, A. & Carter, L. Nanoparticle mediated electron transfer across organic layers: from current understanding to applications. Journal of the Brazilian Chemical Society 25, 418–426 (2014).

20. Elgrishi, N. et al.. A practical beginner’s guide to cyclic voltammetry. Journal of chemical education 95, 197–206 (2018).

21. Hosseini, S. et al.. Synthesis and processing of ELISA polymer substitute: the influence of surface chemistry and morphology on detection sensitivity. Applied surface science 317, 630–638 (2014).

22. Libertino, S. et al.. Layer uniformity in glucose oxidase immobilization on SiO2 surfaces. Applied Surface Science 253, 9116–9123 (2007).

23. Bhattacharyya, S., Cardinaud, C. & Turban, G. Spectroscopic determination of the structure of amorphous nitrogenated carbon films. Journal of applied physics 83, 4491–4500 (1998).

24. Lipińska, W. et al.. The optimization of enzyme immobilization at Au-Ti nanotextured platform and its impact onto the response towards glucose in neutral media. Materials Research Express 6, 1150e1153 (2019).

25. Yang, D. et al.. Chemical analysis of graphene oxide films after heat and chemical treatments by X-ray photoelectron and Micro-Raman spectroscopy. Carbon 47, 145–152 (2009).

26. Xie, W., Abidi, I.H., Luo, Z., Weng, L.-T. & Chan, C.-M. Characterization of the interaction between graphene and copper substrate by time-of-flight secondary ion mass spectrometry. Applied Surface Science 544, 148950 (2021).

27. Wilhelmi, M., Müller, C., Ziegler, C. & Kopnarski, M. BSA adsorption on titanium: ToF-SIMS investigation of the surface coverage as a function of protein concentration and pH-value. Analytical and bioanalytical chemistry 400, 697–701 (2011).

28. Tyler, B., Bruening, C., Rangaranjan, S. & Arlinghaus, H. TOF-SIMS imaging of adsorbed proteins on topographically complex surfaces with Bi3+ primary ions. Biointerphases 6, 135–141 (2011).

29. Eggers, K.M., Jaffe, A.S., Lind, L., Venge, P. & Lindahl, B. Value of cardiac troponin I cutoff concentrations below the 99th percentile for clinical decision-making. Clinical chemistry 55, 85–92 (2009).

30. Mueller, T., Egger, M., Peer, E., Jani, E. & Dieplinger, B. Evaluation of sex-specific cut- off values of high-sensitivity cardiac troponin I and T assays in an emergency department setting–Results from the Linz Troponin (LITROP) study. Clinica Chimica Acta 487, 66–74 (2018).

31. Redfield, M.M. et al.. Plasma brain natriuretic peptide concentration: impact of age and gender. Journal of the American College of Cardiology 40, 976–982 (2002).

32. Ponikowski, P. et al.. 2016 ESC Guidelines for the diagnosis and treatment of acute and chronic heart failure: The Task Force for the diagnosis and treatment of acute and chronic heart failure of the European Society of Cardiology (ESC) Developed with the special contribution of the Heart Failure Association (HFA) of the ESC. European heart journal 37, 2129–2200 (2016).

33. Cevik, S. et al.. NRGN, S100B and GFAP levels are significantly increased in patients with structural lesions resulting from mild traumatic brain injuries. Clinical neurology and neurosurgery 183, 105380 (2019).

