## Supplementary Information for "Rapid antifouling nanocomposite coating enables highly sensitive multiplexed electrochemical detection of myocardial infarction and concussion markers"

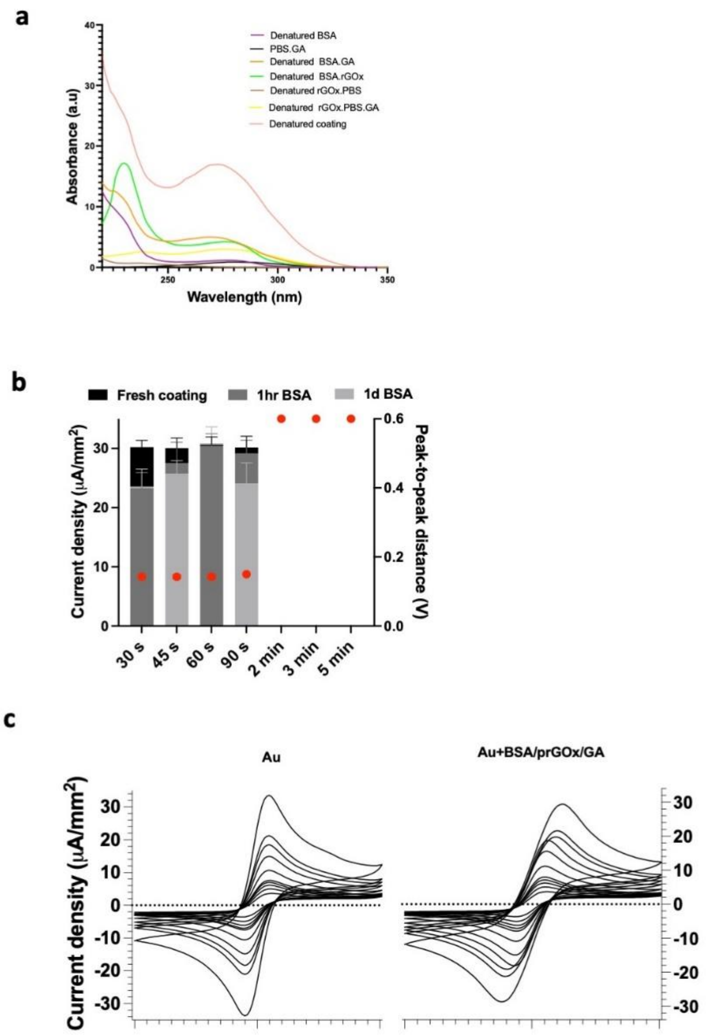


**Supplementary Figure S1.** Characterization of antifouling nanocomposite coating. a, UV absorption spectra of BSA when mixed with/without GA and/or prGOx. a.u., arbitrary units. b, Electrochemical characterization of the coating for development of the optimum heating time where sensors are left at room temperature for 10 min after heating followed by washing in PBS. c, d) Typical voltammograms of bare gold- (left) and coated gold electrodes (right) of an equimolar solution of 5 mM ferri-/ferrocyanide at different scan rates (0.01–1.0 V/s). Error bars represent the s.d. of the mean, n=3.

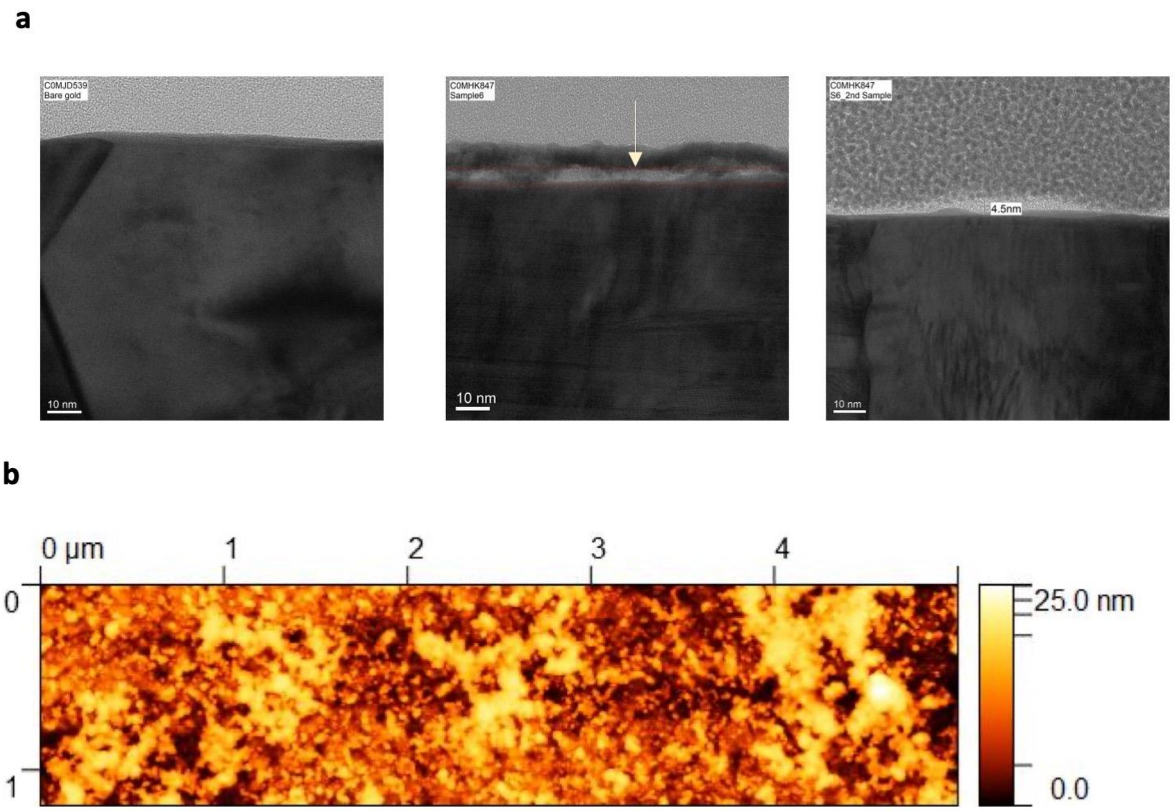


**Supplementary Figure S2.** Characterization of the sensors. a, TEM image of bare gold, antifouling coated gold electrode with top Iridium layer and without top Iridium layer for better contrast. b, AFM image of Gold+BSA/prGOx/GA (anti-fouling coating).

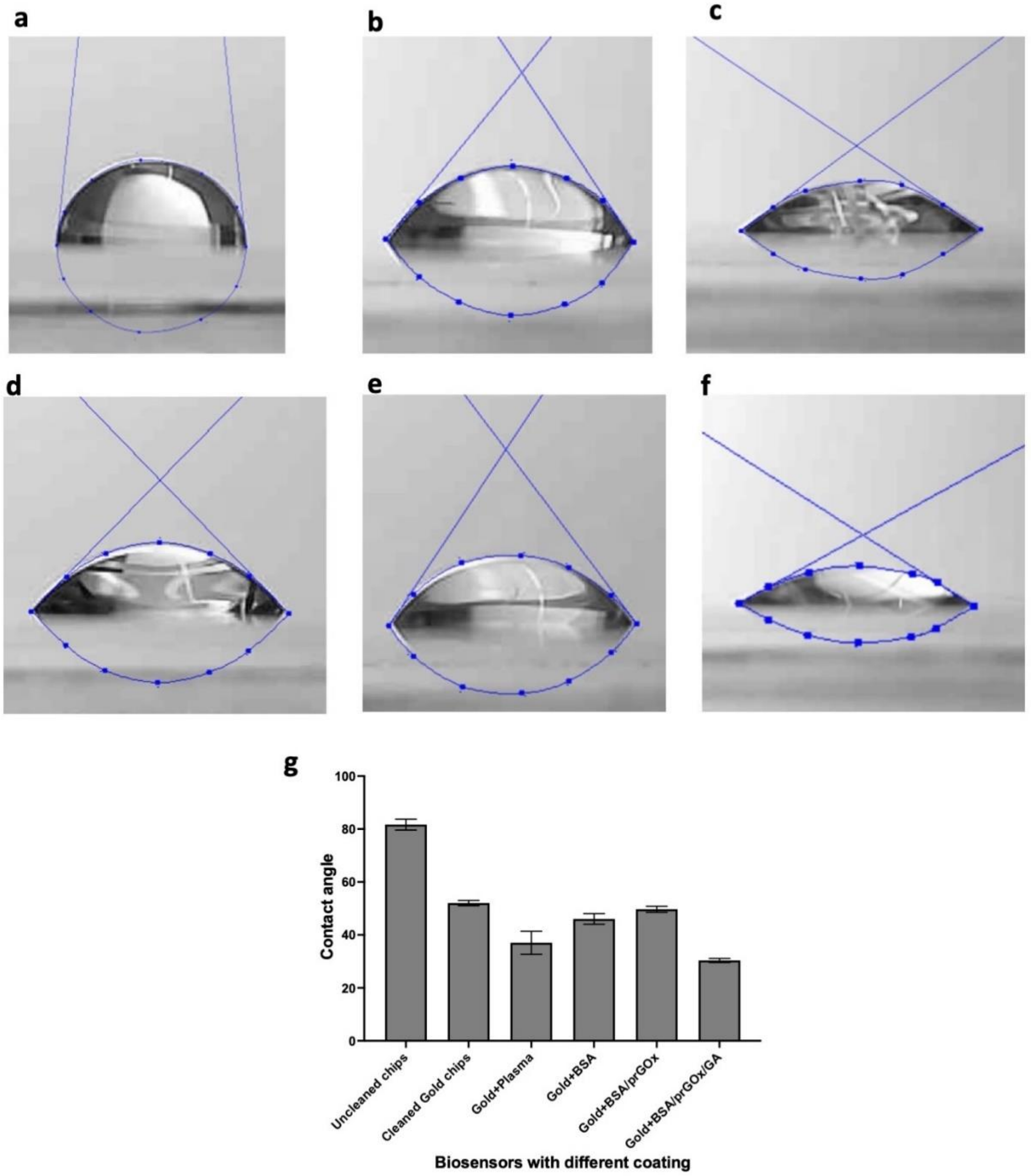


**Supplementary Figure S3.** Measurement of the contact angle of the sensor. a, uncleaned chips with protective organic layer. b, Chips cleaned with acetone and Isopropyl alcohol. c, Plasma treated chips. d, Plasma treated chips with BSA coating. e, Plasma treated chips with BSA/prGOx coating. f, Plasma treated chips with Gold+BSA/prGOx/GA (antifouling nanocomposite coating). g, Contact angle of the EC-Biosensor before and after cleaning and plasma treatment with different kinds of coatings.

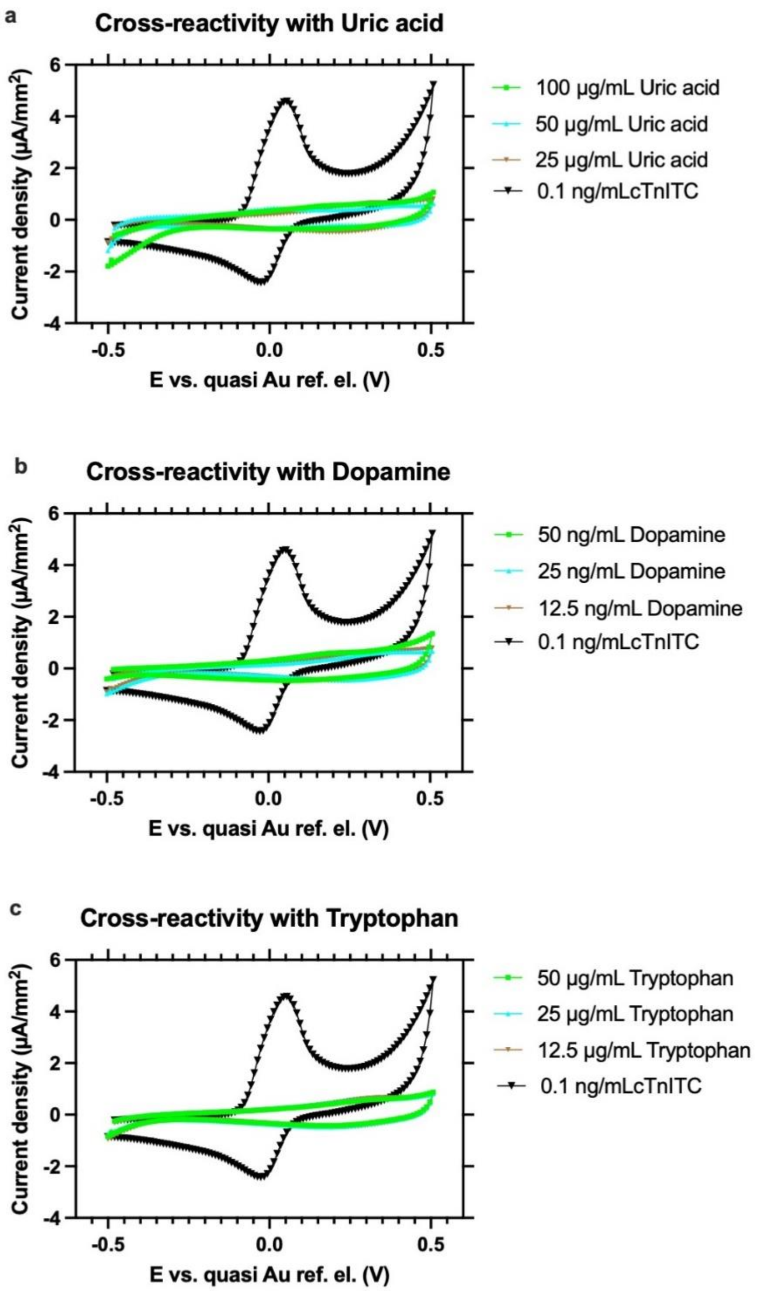


**Supplementary Figure S4.** Cross-reactivity test of the EC Biosensor. Cyclic voltammograms oxidation and reduction peaks of uric acid (a), Dopamine (b), and Tryptophan (c) at 3 different concentrations (at, higher, and lower than physiological level) along with cTnI-TC at 0.1 ng/mL.

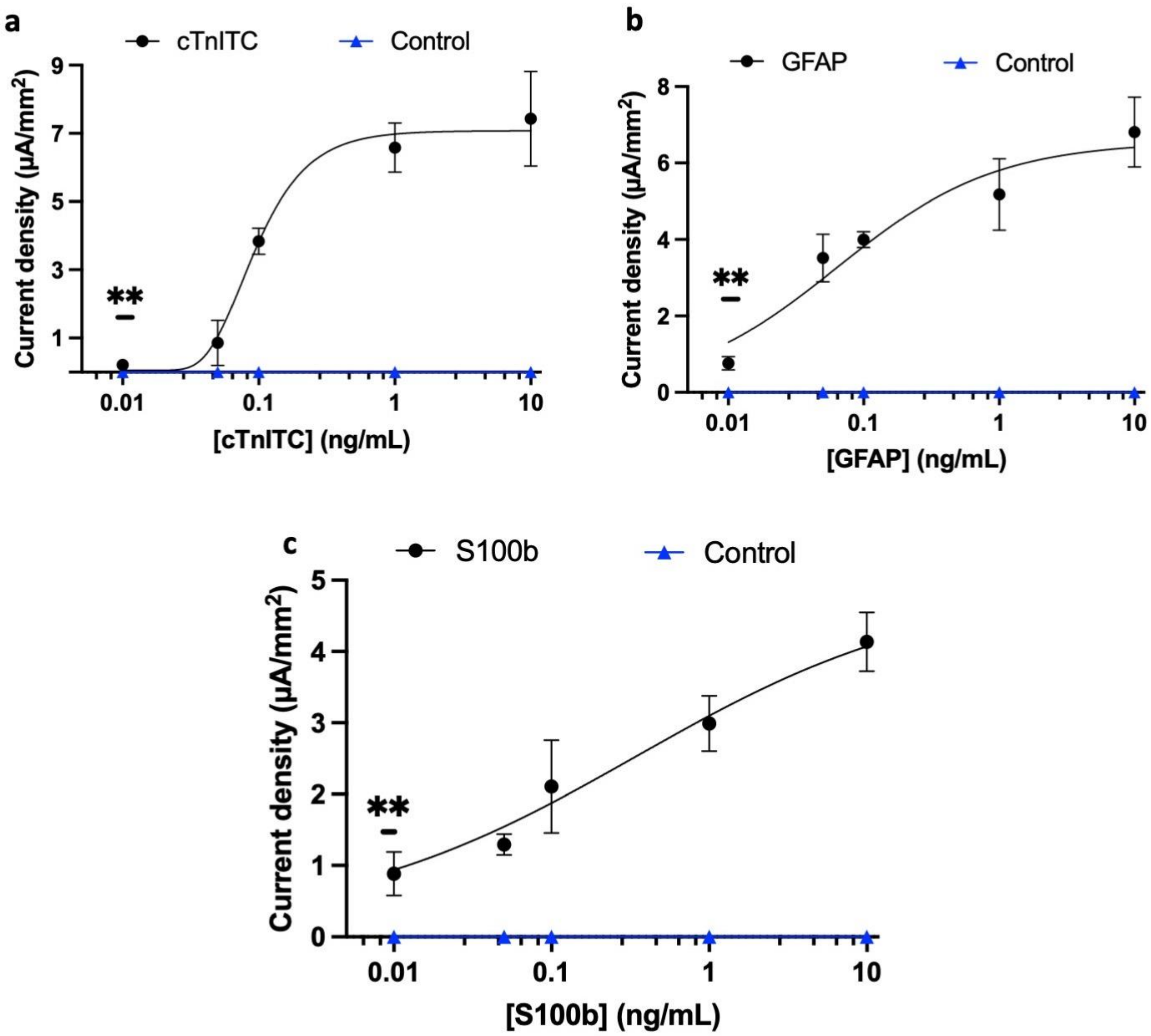


**Supplementary Figure S5.** Calibration curve of different biomarkers run on the EC Biosensor with antifouling coating using unprocessed whole blood. Y-axis shows the current density for different concentrations of biomarkers run on EC Biosensors. a, Calibration curve of cTnITC; b, Calibration curve of GFAP, and c, Calibration curve of S100b. Error bars represent the s.d. of the mean, n=3. Analysis was done using 4-Parameter Logistic (4PL) curve fitting. Lowest concentration showing significant difference against background (0 ng/mL) was determined by unpaired *t*-test ($^{ns}P > 0.05$; $*P < 0.05$; $**P < 0.01$; all two tailed.

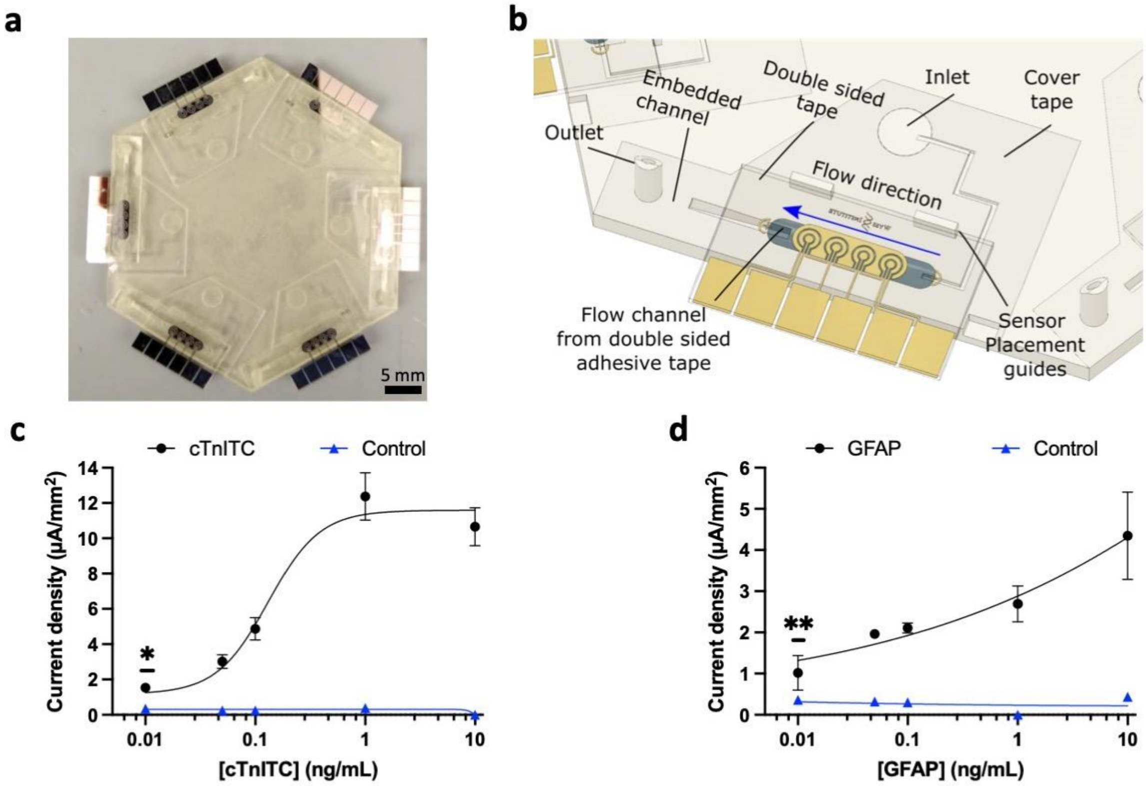


**Supplementary Figure S6.** Microfluidic integration of the assay. a, Microfluidic device where six EC Biosensors can be placed to run the assay in parallel. b, 3D schematic of the microfluidic channels and their interface with the EC Biosensor. c, Calibration curve for the assay of cTnITC performed on the microfluidic platform with reduced assay time using spiked plasma samples. d, Calibration curve for the assay of GFAP performed on the microfluidic platform using spiked plasma samples. Scale bar size: 5 mm. Error bars represent the s.d. of the mean, n=3. In c and d, analysis was done using 4-Parameter Logistic (4PL) curve fitting. Lowest concentration showing significant difference against background (0 ng/mL) was determined by unpaired *t*-test ($^{ns}P > 0.05$; $^{*}P < 0.05$; $^{**}P < 0.01$; both two tailed.

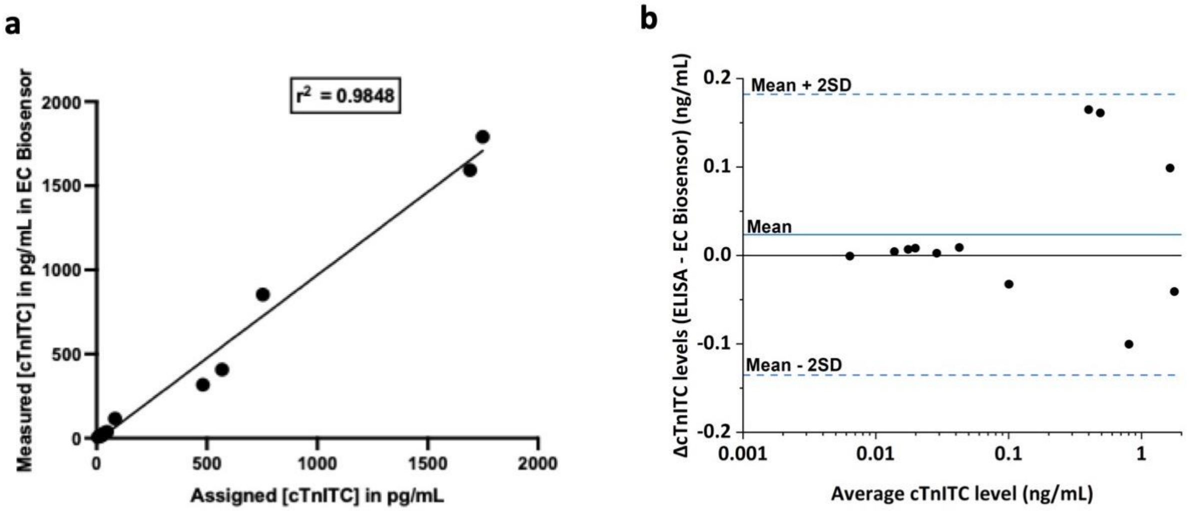


**Supplementary Figure S7**. Clinical validation for cTnI-TC complex on EC-Biosensors. a, Validation of the EC-Biosensor using standard ELISA assay for the detection cTnI-TC complex with n=3 for each clinical sample. b, Bland-Altman plot for validation of EC-Biosensor using clinical sample for cTnI-TC complex. Error bars represent the s.d. of the mean.

**Supplementary Table S1.** Roughness Results for AFM

| Sample | $R_q$ (nm) | $R_a$ (nm) | $R_{max}$ (nm) | Surface Area Diff (%) |
|---|---|---|---|---|
| **Bare Gold** | 2.32 | 1.8 | 22.9 | 2.61 |
| **Coating** | 3.73 | 2.92 | 27.5 | 5.15 |

RMS ($R_q$) is the standard deviation of the Z values (or RMS roughness) in the image. It was calculated according to the formula: $R_q$=Ö{S($Z_i$-$Z_{avg}$)$^2$/N}, where $Z_{avg}$ is the average Z value within the image; $Z_i$ is the current value of Z, and; N is the number of points in the image.

Mean roughness ($R_a$) is the mean value of the surface relative to the Center Plane and was calculated using the formula: $R_a$=[1/($L_x L_y$)]ò$_0^{Ly}$ò$_0^{Lx}${f(x,y)}dxdy, where f(x,y) is the surface relative to the Center Plane, and $L_x$ and $L_y$ are the dimensions of the surface.

Max height ($R_{max}$) is the difference in height between the highest and lowest points of the surface relative to the Mean Plane. Surface area is the area of the 3-dimensional surface of the imaged area. It was calculated by taking the sum of the areas of the triangles formed by 3 adjacent data points throughout the image.

Surface area diff is the amount that the Surface area is over the imaged area. It was expressed as a percentage and is calculated according to the formula: Surface area diff = 100[(Surface area/$S_1^2$)-1], Where $S_1$ is the length (and width) of the scanned area minus any areas excluded by stopbands.

**Supplementary Table S2**. Comparison of sensitivity and Kd for detection of MI and TBI biomarkers.

| Biomarker | Device | Sample | Time | Volume | Kd | Sensitivity |
|---|---|---|---|---|---|---|
| cTnI | EC Biosensor | Plasma | 37 min | 15 µL | 0.5810 | 9 pg/mL |
| | ELISA | Plasma | 2h 40 min | 100 µL | 14.90 | 67 pg/mL |
| BNP | EC Biosensor | Plasma | 37 min | 15 µL | 0.190 | 26 pg/mL |
| | ELISA | Plasma | 2h 40 min | 100 µL | ~ 187331 | 2783 pg/mL |
| NT-proBNP | EC Biosensor | Plasma | 37 min | 15 µL | 1.323 | 4 pg/mL |
| | ELISA | Plasma | 2h 40 min | 100 µL | 3.509 | 65 pg/mL |
| cTnITC | EC Biosensor | Plasma | 37 min | 15 µL | 0.1663 | 3 pg/mL |
| | EC Biosensor (microfluidics) | Plasma | 15 min | 40 µL | 0.083 | 14 pg/mL |
| | EC Biosensor | Whole blood | 37 min | 15 µL | 0.0837 | 22 pg/mL |
| | ELISA | Plasma | 2h 40 min | 100 µL | 1.190 | 24 pg/mL |
| GFAP | EC Biosensor | Plasma | 37 min | 15 µL | 0.0688 | 5 pg/mL |
| | EC Biosensor (microfluidics) | Plasma | 15 min | 40 µL | - | 27 pg/mL |
| | EC Biosensor | Whole blood | 37 min | 15 µL | 0.0642 | 2 pg/mL |
| | ELISA | Plasma | 2h 40 min | 100 µL | 83.59 | 59 pg/mL |
| S100b | EC Biosensor | Plasma | 37 min | 15 µL | 6.105 | 1 pg/mL |
| | EC Biosensor | Whole Blood | 37 min | 15 µL | 0.3403 | 13 pg/mL |
| | ELISA | Plasma | 2h 40 min | 100 µL | ~ 1.0e+016 | 957 pg/mL |
